# Model the transmission dynamics of COVID-19 propagation with public health intervention

**DOI:** 10.1101/2020.04.22.20075184

**Authors:** Dejen Ketema Mamo

## Abstract

In this work, a researcher develop *SHEIQRD* (Susceptible-Stay at home-Exposed-Infected-Quarantine-Recovery-Death) coronavirus pandemic spread model. The disease-free and endemic equilibrium points are calculated and analyzed. The basic reproductive number *ℛ*_0_ is derived and its sensitivity analysis is done. COVID-19 pandemic spread is die out when *ℛ*_0_ ≤ 1 and its persist in the community whenever *ℛ*_0_ > 1. Efficient stay at home rate, high coverage of precise identification and isolation of expose and infected individuals, and redaction of transmission and stay at home return rate can be mitigate the pandemics. Finally, theoretical analysis and numerical results are consistent.

## 1. Introduction

Coronaviruses are a large family of viruses which may cause illness in animals or humans. In humans, several coronaviruses are known to cause respiratory infections ranging from the common cold to more severe diseases such as Middle East Respiratory Syndrome (MERS) and Severe Acute Respiratory Syndrome (SARS). The most recently discovered coronavirus causes Coronavirus disease 2019(COVID-19) [1]. Its the infectious disease caused by the most recently discovered coronavirus. This new virus and disease were unknown before the outbreak began in Wuhan, China, in December 2019. The most common symptoms of COVID-19 are fever, tiredness, and dry cough. Some patients may have aches and pains, nasal congestion, runny nose, sore throat or diarrhea. These symptoms may appear 2-14 days after exposure, most commonly around five days [2, 3].

China was the index case of COVID-19 pandemic later it rapidly spread thought the world. People infected by those initial cases spread the disease to other drastically due to human to human transmission [4]. Although Corona represents a major public health issue in world, as of March 11, 2020, over 118,000 infections spanning 113 countries have been confirmed by the World Health Organization (WHO). The WHO declared this public health emergency as a pandemic [5]. As of 14 April 2020, WHO reported 1, 844, 863 confirmed case and 117, 021 deaths have been recorded globally [6].

The study about the spread and control of COVID-19 is essential at this time. Different scholars are study about infectious disease spread control by using modeling approach [7, 8, 9, 10, 11, 12]. Recently, researcher study a bout COVID-19 [13, 14, 15, 16]. The model, which is of *SEIR* form [17], incorporates the recommended public health interventions in the current pandemic. The recommended mitigation strategies of the pandemic are stay at home, and isolation of expose and infected individuals by efficient identification process. A researcher focus on the impact of control measures by varying the parameter values. The model result indicates that the containment of the pandemic requires high level of both identification and isolation process and the contact tracing process by stay at home for removing infected individuals from the susceptible population.

## 2. Model Formulation

In this work a researcher consider that the total population is *N* (*t*) at time *t*. The whole population dividing in to seven compartments. The susceptible population *S*(*t*), they stand for people who are capable of becoming infected. The quarantine population *H*(*t*), they represent people who are stay at home. The exposed population *E*(*t*), they represent people who are incubating the infection. The spreader population *I*(*t*), they represent people who are infectious infected. The quarantine population *Q*(*t*), they represent people who are isolated by clinically confirmation. The recovery population *ℛ*(*t*), they represent people who are discharge from the virus. The density of disease induced death denoted by *D*(*t*).

The model flows chart is describes in figure 1.

**Figure 1:**
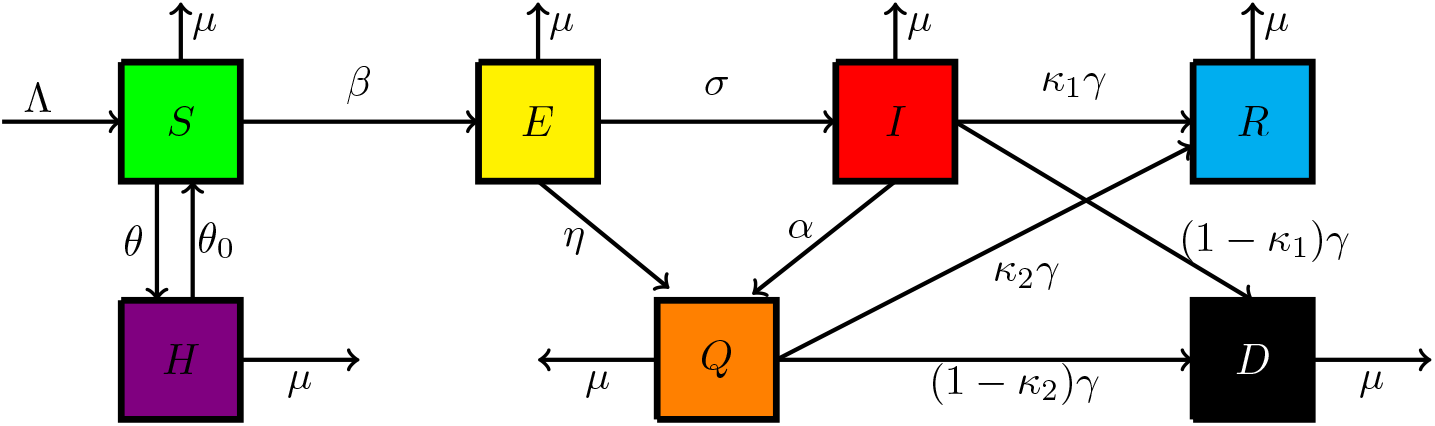
Schematic diagram of the model compartments and parameters.

In the process of COVID-19 spreading, the spreading among these seven states is governed by the following assumptions. It is assumed that *β* is the contact rate of susceptible individuals with spreaders and the disease transmission follows the mass action principle. A researcher assume that susceptible individuals home quarantine or stay at home at the rate *θ*. And at a rate *θ*_0_ staying at home is not fully protected from the virus due to ineffectiveness of home quarantine. The one who completed incubation period becomes to infected at a rate of *σ*, that means 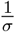 is the average duration of incubation. According to clinical examination, the exposed and infected individuals becomes isolated at a rate of *η* and *α* respectively. It is assumed that the infectious infected individuals, leading to disease prevalence. The average duration of infectiousness is 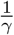, when *γ* is the transmission rate from infected to recovery or death. In my assumption recovery from isolated infected is better than and infected class due to clinical treatment. Infected and isolated infected are recover with a probability of *κ*_1_and *κ*_2_, and also they will becomes to death with a probability of (1 − *κ*_1_) and (1 − *κ*_2_) respectively. The parameter Λ is the recruitment, while *µ* natural birth and death rate of each state individuals. The parameters are all non-negative. Based on the above considerations, COVID-19 spreading leads to dynamic transitions among these states, shown in figure 1. The model can be described by the following system of nonlinear ordinary differential equations:

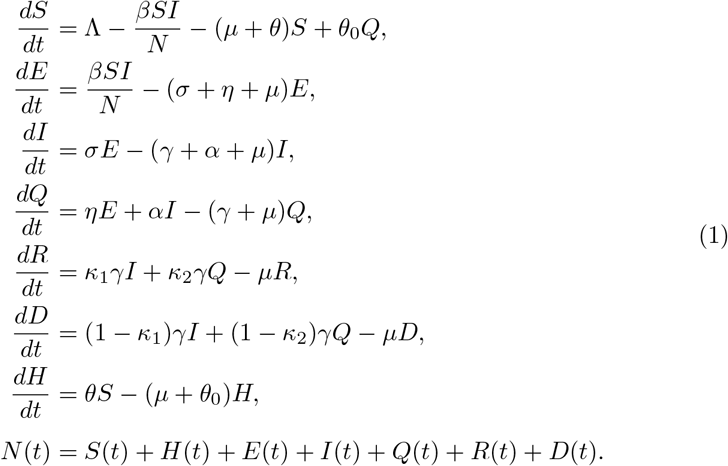

We have the non-negative initial conditions 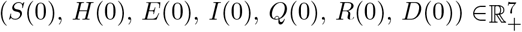

To make the mathematical analysis more easier, the variables of the model (1)can be normalized as 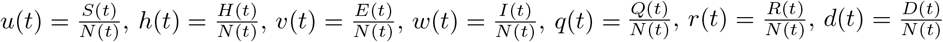, and Λ = *µN* (*t*). After substitute it in (1) we can get the simplified form of the model

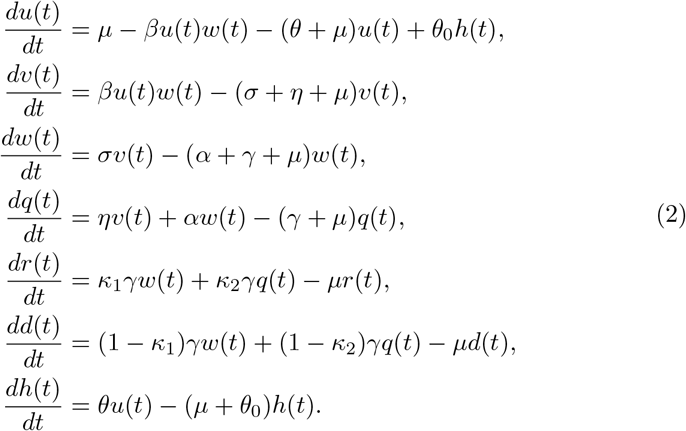

From the normalized form of the model we have to get

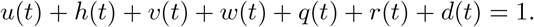

Now, the first equation of the system (2) can be reduced and we hold six system of differential equations

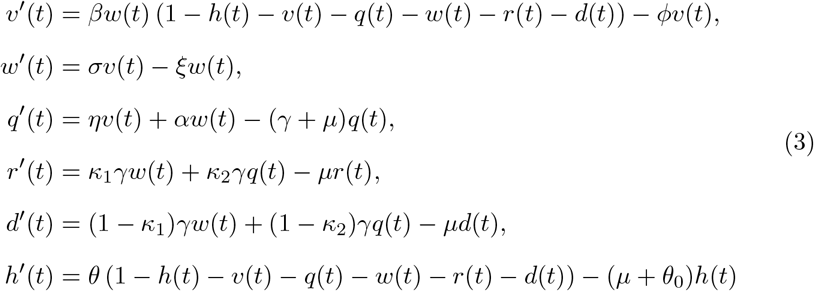

where *φ* = (*σ* + *η* + *µ*) and *ξ* = (*α* + *γ* + *µ*).

So, the feasible domain of the system (3) is

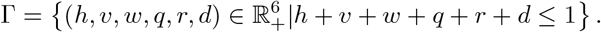

For the well-posedness of the model, we have the following lemma.

### Lemma 1.

*The set* Γ *is positively invariant to system* (3).

Proof. Denote *x*(*t*) = (*h*(*t*), *v*(*t*), *w*(*t*), *q*(*t*), *r*(*t*), *d*(*t*))^*T*^ and then system (3) can be rewritten as

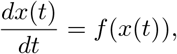

Where

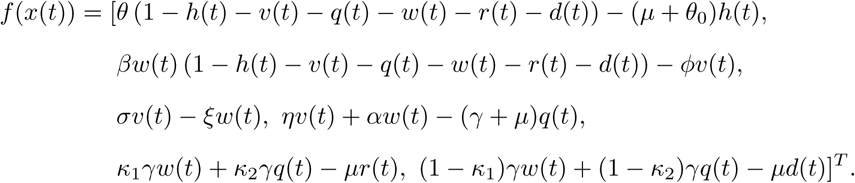

Note that Ω is obviously a compact set. We only need to prove that if *x*(0) *∈* Γ, then *x*(*t*) *∈* Γ for all *t ≥* 0. Note that *∂*Γ consists of five plane segments:

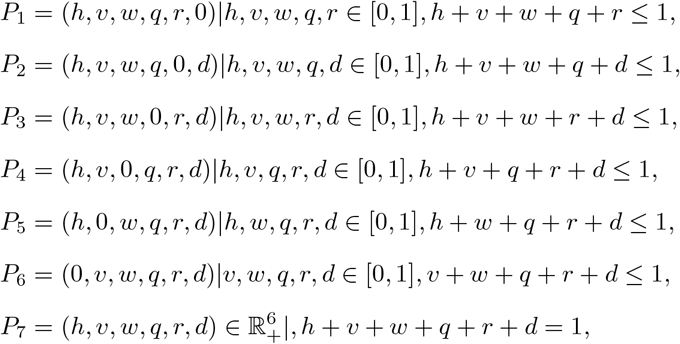

which have *v*_1_ = (0, 0, 0, 0, 0, −1), *v*_2_ = (0, 0, 0, 0, −1, 0), *v*_3_ = (0, 0, 0, −1, 0, 0), *v*_4_ = (0, 0, −1, 0, 0, 0), *v*_5_ = (0, −1, 0, 0, 0, 0), *v*_6_ = (0, −1, 0, 0, 0, 0, 0), *v*_7_ = (1, 1, 1, 1, 1, 1) as their outer normal vectors, respectively. If the dot product of *f* (*x*) and normal vectors (*v*_1_, *v*_2_, *v*_3_, *v*_4_, *v*_5_, *v*_6_, *v*_7_) of the boundary lines are less than or equal to zero, then *x*(*t*) *∈* Γ for all *t ≥* 0. So,

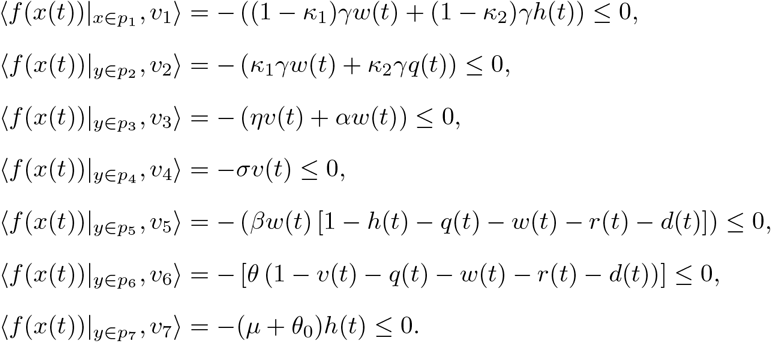

The proof is complete.

Hence, system (1) is considered mathematically and biologically well posed in Γ[18].

## 3. Theoretical analysis of the model

### 3.1. Equilibrium analysis

In this sub section, we show the feasibility of all equilibria by setting the rate of change with respect to time *t* of all dynamical variables to zero. The model (2) has two feasible equilibria, which are listed as follows:

(i) Disease-free equilibrium (DFE)*E*_0_ 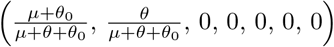

(ii) Endemic equilibrium (EE) *E*^*\**^ (*u*^*\**^, *h*^*\**^, *v*^*\**^, *w*^*\**^, *q*^*\**^, *r*^*\**^, *d*^*\**^).

The existence of endemic equilibrium is computed after we have the basic reproductive number *ℛ*_0_.

### 3.2. Basic reproduction number

Here, we will find the basic reproduction number (*ℛ*_0_) of the model (2) using next generation matrix approach [19]. We have the matrix of new infection *F*(*X*) and the matrix of transfer *V*(*X*). Let *X* = (*v, w, q, h, u, r, d*)*T*, the model (2) can be rewritten as:

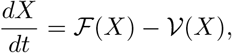

Where

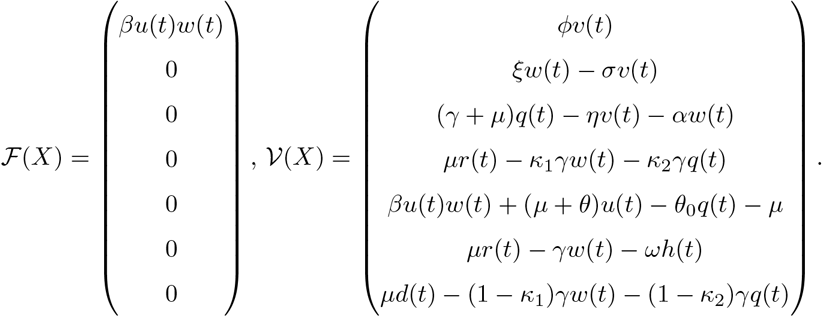

The Jacobian matrices of *ℱ*(*X*) and *𝒱*(*X*) at the disease free equilibrium 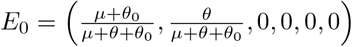 are, respectively,

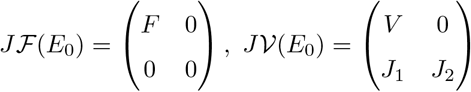

where,

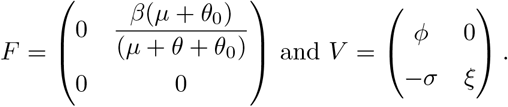

The inverse of *V* is computed as

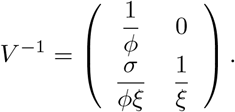

The next generation matrix *𝒦*_*L*_ = *FV* ^−1^ is given by

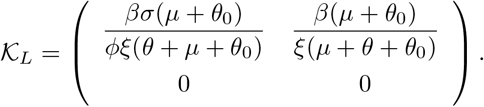

Therefore, basic reproduction number is *ℛ*_0_ = *ρ*(*𝒦*_*L*_) = max (|*µ*| : *µ ∈ ρ*(*𝒦*_*L*_)) is spectral radius of matrix *𝒦*_*L*_ and basic reproduction number (*ℛ*_0_) is obtained as follows,

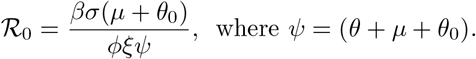

### 3.3. Stability of the disease free equilibrium

In this subsection, we summarize the results of linear stability of the model by finding the sign of eigenvalues of the Jacobian matrix around the equilibrium *E*_0_.

#### Theorem 2.

*If ℛ*_0_ *<* 1, *the disease-free equilibrium E*_0_ *of system* (2) *is locally asymptotically stable, and it is unstable if ℛ*_0_ > 1.

Proof. In the absence of the disease, the model has a unique disease free equilibrium *E*_0_. Now the Jacobian matrix at equilibrium *E*_0_ is given by:

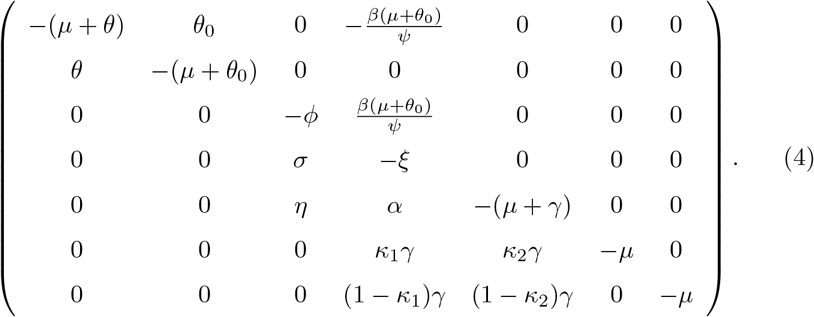

Here, we need find the eigenvalue of the system from the Jacobian matrix (4). We obtain the characteristic polynomial

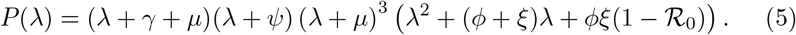

From the characteristic polynomial in equation (5), it is easy to get five real negative eigenvalues of *J* (*E*_0_), which are *λ*_1,2,3_ = −*µ, λ*_4_ = −*µ*−*γ* and *λ*_5_ = −*ψ*. We get the other real negative eigenvalues from the expression

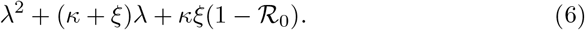

From the quadratics equation (6), we conclude that *λ*_6,7_ are positive if *ℛ*_0_ > 1 and negative if *ℛ*_0_ *<* 1. Thus, the equilibrium *E*_0_ is locally asymptotically stable if *ℛ*_0_ *<* 1. *E*_0_ becomes unstable whenever *E*^*\**^ is feasible (i.e., *ℛ*_0_ > 1). The proof is complete.

Physically speaking, theorem 2 implies that disease can be eliminated if the initial sizes are in the basin of attraction of the DFE *E*_0_. Thus the infected population can be effectively controlled if *ℛ*_0_ *<* 1. To ensure that the effective control of the infected population is independent of the initial size of the human population, a global asymptotic stability result must be established for the DFE.

#### Theorem 3.

*If ℛ*_0_ ≤ 1, *then the disease-free equilibrium, E*_0_, *of system* (2) *is globally asymptotically stable in* Γ.

Proof. Let *X* = (*u, h, v, w, q, r, d*)^*T*^ and consider a Lyapunov function,

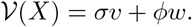

Differentiating *𝒱* in the solutions of system (2) we get

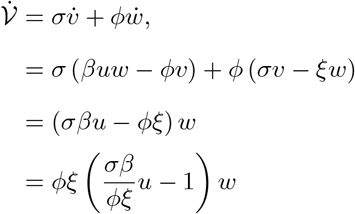

Therefore,

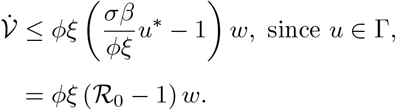

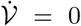 whenever *ℛ*_0_ *<* 1. Furthermore, 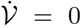 = 0 if and only if *ℛ*_0_ = 1. Thus the largest invariant set in 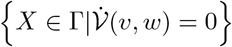 is the singleton, 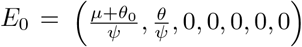. By LaSalle’s Invariance Principle the disease-free equilibrium is globally asymptotically stable in Γ, completing the proof.

Theorem 3 completely determines the global dynamics of model (2) in when *ℛ*_0_ ≤ 1. It establishes the basic reproduction number *ℛ*_0_ as a sharp threshold parameter. Namely, if *ℛ*_0_ *<* 1, all solutions in the feasible region converge to the DFE *E*_0_, and the disease will die out from the community irrespective of the initial conditions. If *ℛ*_0_ > 1, *E*_0_ is unstable and the system is uniformly persistent, and a disease spread will always exist.

### 3.4. Endemic equilibrium and its stability

#### 3.4.1. Existence and uniqueness

The feasibility of the equilibrium *E*_0_ is trivial. Here, we show the feasibility of endemic equilibrium *E*^*\**^. The values of *u*^*\**^, *h*^*\**^, *v*^*\**^, *w*^*\**^, *q*^*\**^, *r*^*\**^ and *d*^*\**^ are obtained by solving following set of algebraic equations:

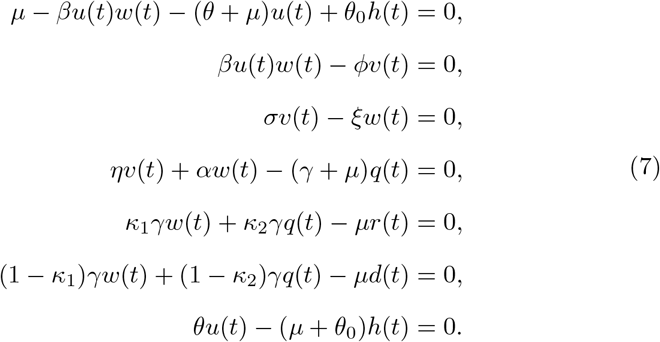

After some algebraic calculations we get the value of *E*^*\**^ as:

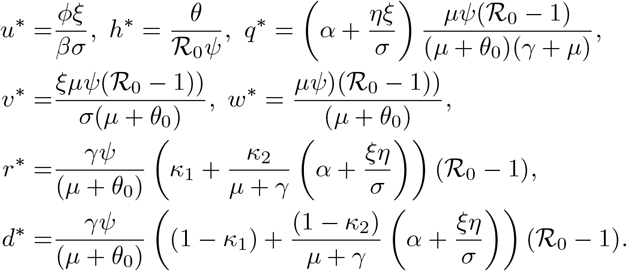

Therefore, there exists a unique positive solution only when *ℛ*_0_ > 1. This implies that, it has a unique endemic equilibrium, *E*^*\**^.

#### 3.4.2. Stability analysis

##### Theorem 4.

*If ℛ*_0_ > 1, *then the endemic equilibrium point E*^*\**^ *of system* (2) *is locally asymptotically stable*.

Proof. The Jacobian matrix of the model at *E*^*\**^ is

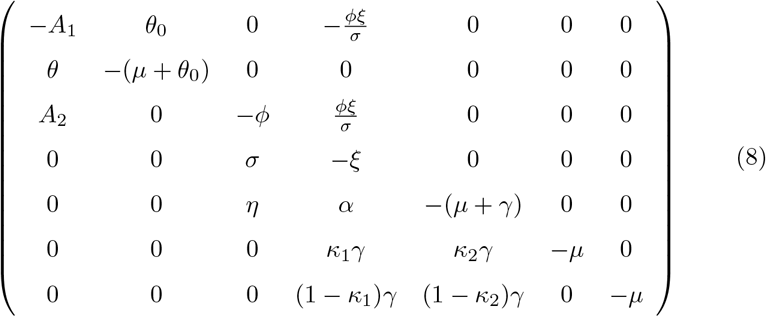

where 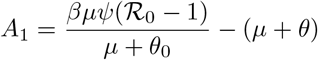 and *A* = *A* + (*µ* + *θ*).

From the Jacobian matrix (8) easily to get *λ*_1,2_ = −*µ, λ*_3_ = −*µ* − *γ* and the other eigenvalues of the system needs further finding. The characteristic polynomial of (8) is

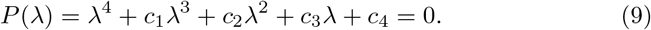

Where

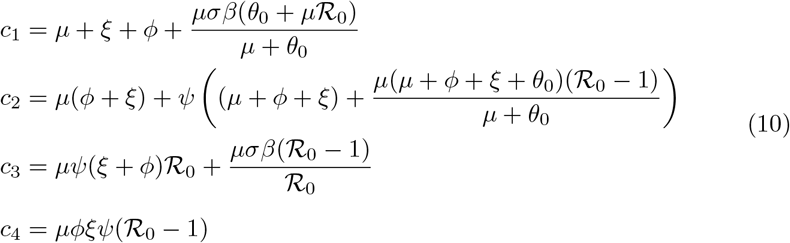

The polynomial (9) has negative roots (eigenvalues) if all its coefficients terms are positive, or it satisfies Routh-Hurwitz criteria of stability [20]. From (10) we can verify that *c*_1_ > 0, *c*_4_ > 0, *c*_1_*c*_2_ − *c*_3_ > 0 and 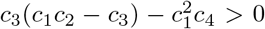, when *ℛ*_0_ > 1. Therefore, according to the Routh-Hurwitz criterion, we can get that all the roots of the above characteristic equation have negative real parts. Thus, the endemic equilibrium asymptotically stable. The proof is complete.

The local stability analysis of the endemic equilibrium tells that if the initial values of any trajectory are near the equilibrium *E*^*\**^, the solution trajectories approach to the equilibrium *E*^*\**^ under the condition *ℛ*_0_ > 1. Thus, the initial values of the state variables *u, h, v, w, q, r* and *d* are near to the corresponding equilibrium levels, the equilibrium number of infected individuals get stabilized if *ℛ*_0_ > 1.

### 3.5. Sensitivity analysis of ℛ_0_

We explore *ℛ*_0_ sensitivity analysis of system (2) to determine the model robustness to parameter values. This is a strategy to identify the most significance parameters of the model dynamics. The normalized sensitivity index Υ_*λ*_ is given by

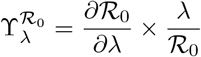

Thus normalized sensitivity indices for parameters are obtained as

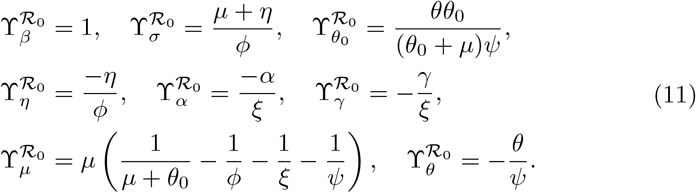

From the sensitivity indices calculation results, we can identify some parameters that strongly influence the dynamics of disease spread. Parameters *β, θ*_0_ and *σ* have a positive influence on the basic reproduction number *ℛ*_0_, that is, an increase in these parameters implies an increase in *ℛ*_0_. While parameters *µ, η, α, θ* and *γ* have a negative influence on the basic reproduction number *ℛ*_0_, that is, an increase in these parameters implies a decrease in *ℛ*_0_.

Here, we illustrate graphically the relationship between the basic reproductive number and the parameters in model (2).

A researcher can find some interesting results, which have been showed in figure 2, and figure 3, it can be seen that big *β* or *σ* can lead to large *ℛ*_0_. That is to say, the larger contact or short incubation period can increase the opportunity of disease spreading. If we reduce the transmission rate by quarantine or any appropriate control measure, then the disease outbreak will end.

**Figure 2:**
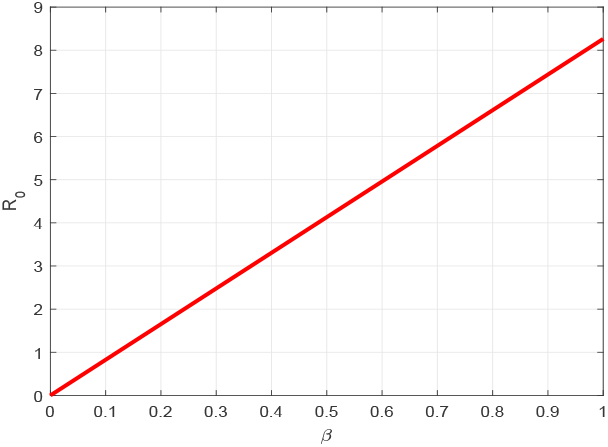
*ℛ*_0_ vs the parameter *β*.

**Figure 3:**
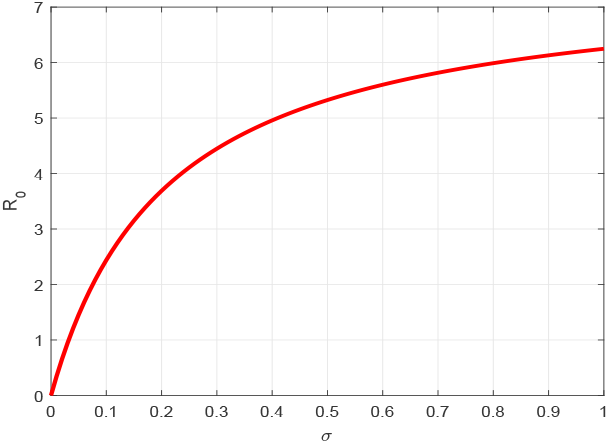
*ℛ*_0_ vs the parameter *σ*.

As a result of figure 4, and figure 5, *ℛ*_0_ decreasing when *θ* increases, and increases whenever *θ*_0_ increase respectively. This finding suggested that effective stay at home intervention have been mitigates the COVID-19 spread, conversely the ineffectiveness of this intervention measure can rising its spread.

**Figure 4:**
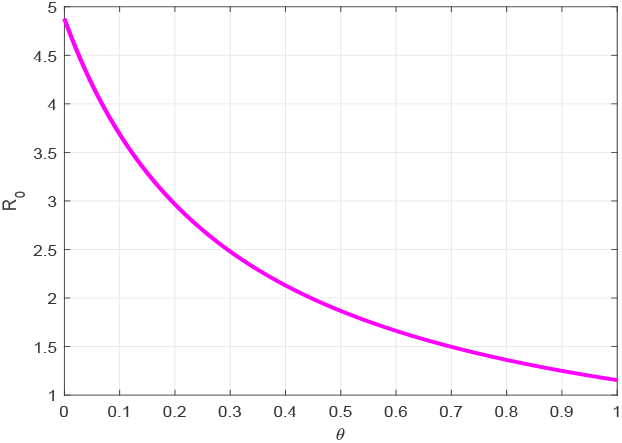
*ℛ*_0_ vs the parameter *θ*.

**Figure 5:**
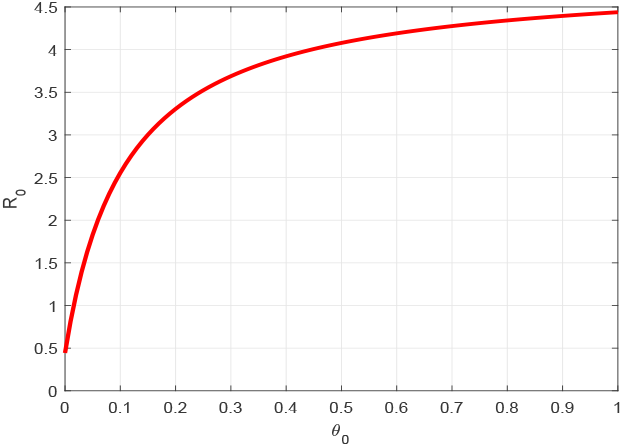
*ℛ*_0_ vs the parameter *θ*_0_.

Figure 6, and figure 7, shows that the increment of *η* or *α* can reduce *ℛ*_0_. That is to say, effective quarantine of incubated and infectious individuals can reduce the opportunity of disease spreading.

**Figure 6:**
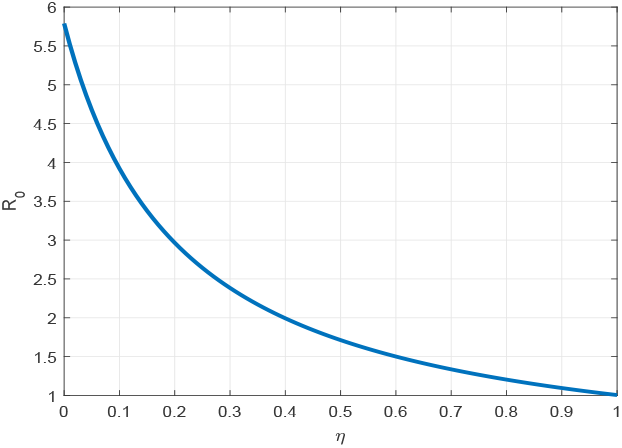
*ℛ*_0_ vs the parameter *η*.

**Figure 7:**
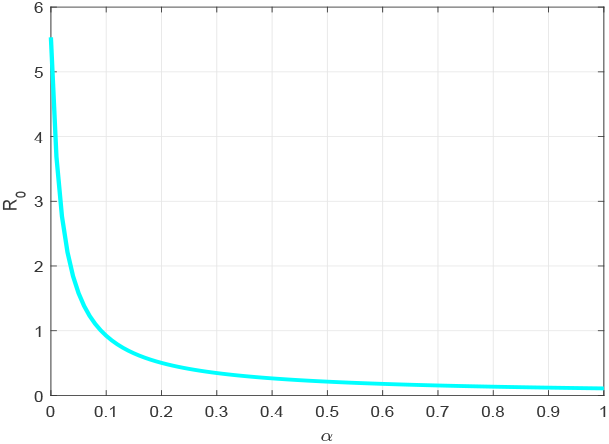
*ℛ*_0_ vs the parameter *α*.

From figure 8, and figure 9, we find that, short average time from the symptom onset to recovery or death *γ* and large value of *µ* can reduce the COVID-19 spread.

**Figure 8:**
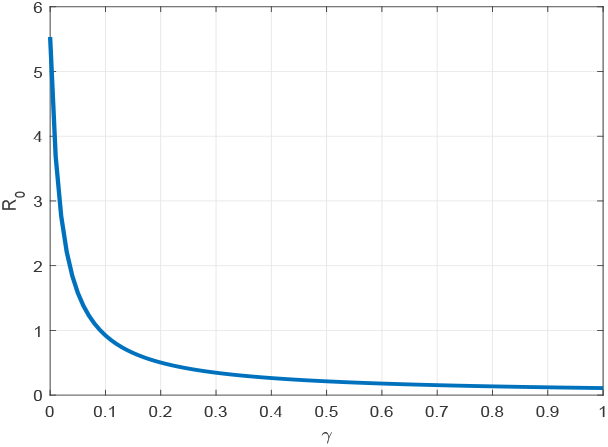
*ℛ*_0_ vs the parameter *γ*.

**Figure 9:**
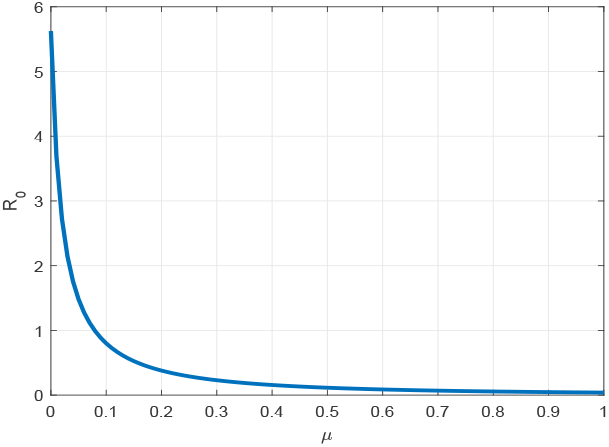
*ℛ*_0_ vs the parameter *µ*.

## 4. Numerical results and analysis

In this section, we conduct numerical simulation of the model (2) by using Matlab standard ordinary differential equations (ODEs) solver function ode45.

### 4.1. General dynamics

We numerically illustrate the asymptotic behavior of the model (2). We take the the initial conditions *u*(0) = 0.9, *q*(0) = 0, *v*(0) = 0.06, *w*(0) = 0.04, *h*(0) = 0, *r*(0) = 0, and *d*(0) = 0.

Figure 10 presents the trajectories of model (2) when *β* = 0.05, *θ* = 0, *σ* = 0.1923, *α* = 0, *γ* = 0.0714, *µ* = 0.01, *θ*_0_ = 0.0, thus the basic reproduction number *ℛ*_0_ = 0.5842. From this figure, we can see that the disease die out and the trajectories converge to the racism free equilibrium point (1, 0, 0, 0, 0, 0, 0). This mean that disease disappears in the community as shown in theorem 2, and theorem 3. Furthermore, socio-economical crisis caused by COVID-19 are removed. Finally, we have to get disease free community.

**Figure 10:**
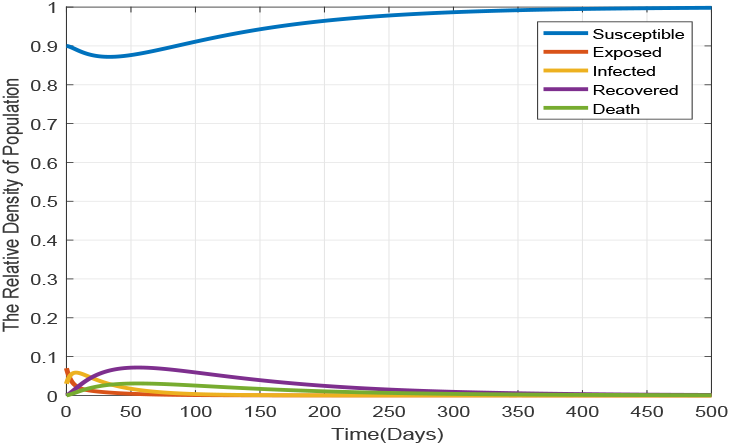
Each compartment population changes over time when *ℛ*_0_ *<* 1.

Figure 11 gives the trajectory plot when *β* = 0.3, *θ* = 0, *σ* = 0.1923, *α* = 0, *γ* = 0.0714, *µ* = 0.01, *θ*_0_ = 0.0, the basic reproduction number is *ℛ*_0_ = 3.5054. From this figure, we can see that even for a small fraction of the infectious case at the beginning, the disease is persists in the community and stabilize in time. This means that the trajectories converge to the endemic equilibrium point. Thus, as established in theorem 4, the disease persists in the community whenever *ℛ*_0_ > 1.

**Figure 11:**
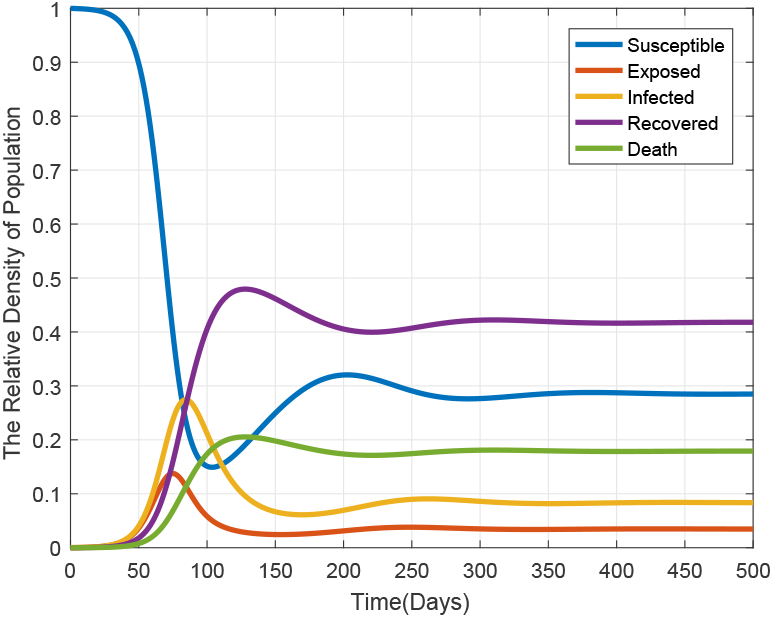
Each compartment population changes over time when *ℛ*_0_ > 1.

### 4.2. Impact of the transmission rate

In order to investigate the impact of the transmission rate on the spread of COVID-19, we carry out a numerical simulation to show the contribution of transmission rate *β* in fractional infection population density.

We set the transmission rate *β* as 0.05, 0.2, 0.25, 0.35 and *β* = 0.5. From figure 12, we can observe that infectiousness reach a higher peak level as *β* increases. This figure illustrates the great influence of transmission rate as shown in the sensitivity analysis. The transmission rate and the basic reproductive number are almost symmetrical relationship (i.e *ℛ*_0_ = 10*×β*). It says that, if we implemented effective contact tracing process between infected and susceptible population, then the transmission rate is reduced and also the disease spread will be eliminated. The main public health measure which are implemented to reduce the transmission rate in the current pandemic are stay at home and quarantine or isolation of exposed and infected individuals by clinical tests.

**Figure 12:**
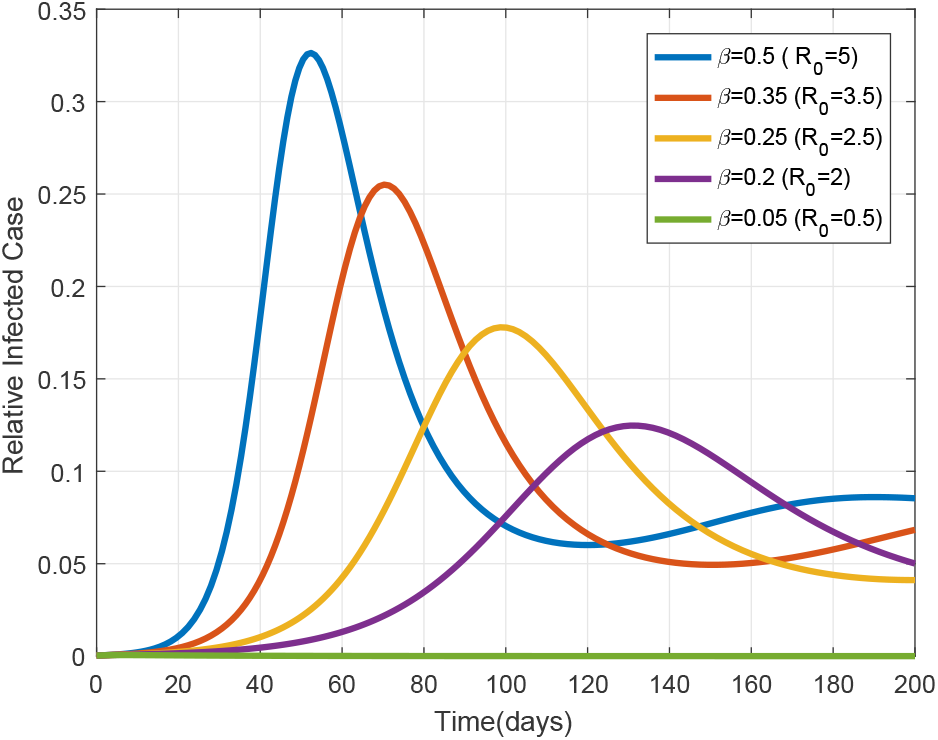
Impact of transmission rate *β* on infected population *w*(*t*) in system (2). Colors represent different values of *β*.

### 4.3. Impact of public health intervention

Study the recommended containment strategies of the pandemic, we conduct some numerical simulations to show the contribution of public health interventions.

One of the recommended control measure to reduce the pandemic is quarantine or isolation. Here, we observe the isolation of exposed and infected individuals within different rate:

Now, we set the exposed population isolation rates *η* as 0.6, 0.2, 0.1, 0.05 and In figure 13, we can see that the infectiousness increase as *η* decrease. This implies that effective isolation of exposed individuals by clinical identification before the symptom onset can mitigates the COVID-19 pandemic. Similarly, infected isolation rates *α* set as 0.25, 0.1, 0.05, 0.03 and 0.0. In figure 14, we observe that the infectiousness density approaches to highest peak level as *α* value decreases. This implies that ineffective quarantine of symptomatic individuals can lead the prevalence of the pandemic.

**Figure 13:**
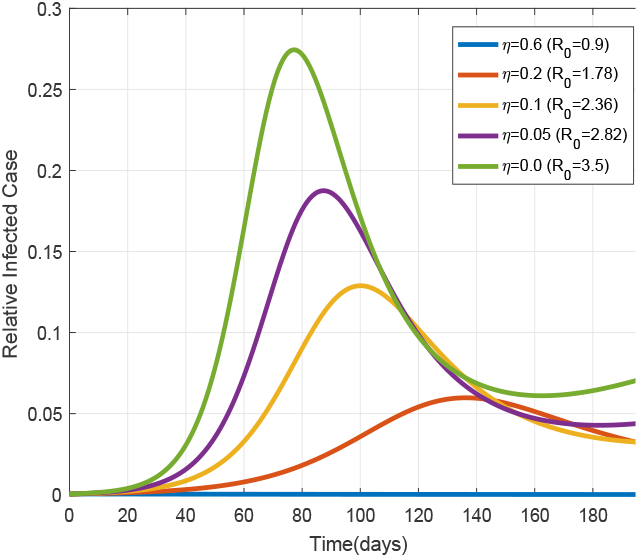
Impacts of *η* on *w*(*t*).

**Figure 14:**
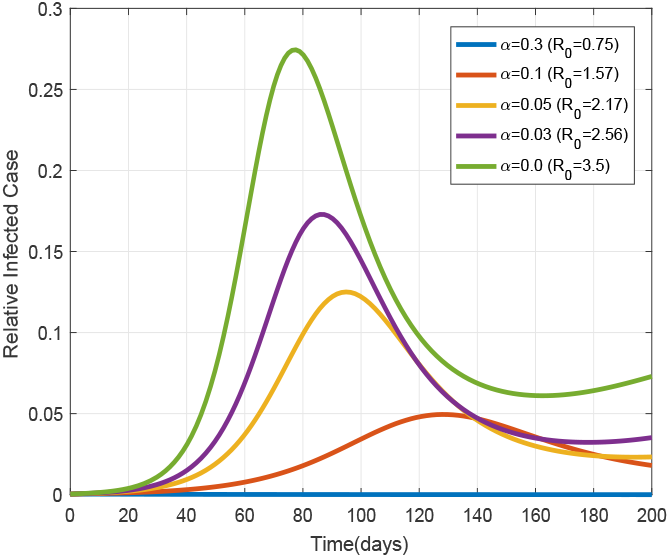
Impacts of *α* on *w*(*t*).

In the current critical time the public health experts and government officials announced that every individuals must stay at home. Due to food security and ineffectiveness of stay at home peoples my lose this recommendation. To observe the impact of stay at home efficiency and its lose in the following numerical results.

Figure 15, shows that different stay home rates *θ*, which are chosen as 0.1, 0.015, 0.01, 0.004 and 0.0. Its say that effective stay at home intervention measure can be control the disease propagation. In the other hand if we can’t implement effectively this control measure, then people becomes to susceptible at a rate of *θ*_0_. To show its impact with *θ* = 0.1, we chose different *θ*_0_ values as 0.6, 0.24, 0.013, 0.0065 and 0.0. We can be see in figure 16, the disease spread rises as *θ*_0_ values increases. This implies if we can’t stay at home with a recommended time span, then we lead the pandemic prevalence.

**Figure 15:**
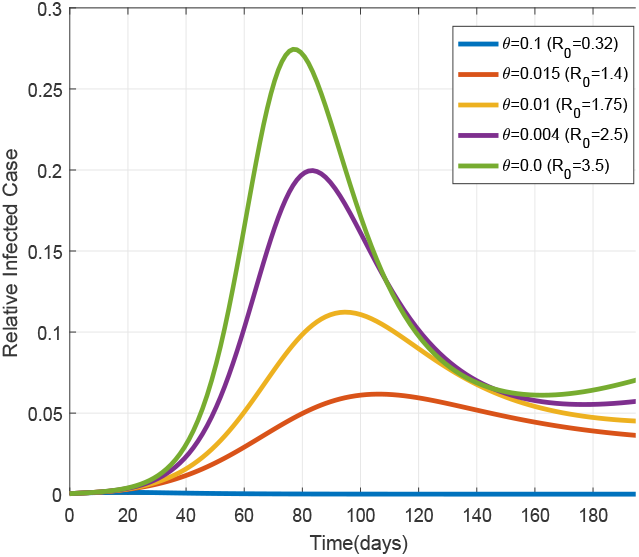
Impacts of *θ* on *w*(*t*).

**Figure 16:**
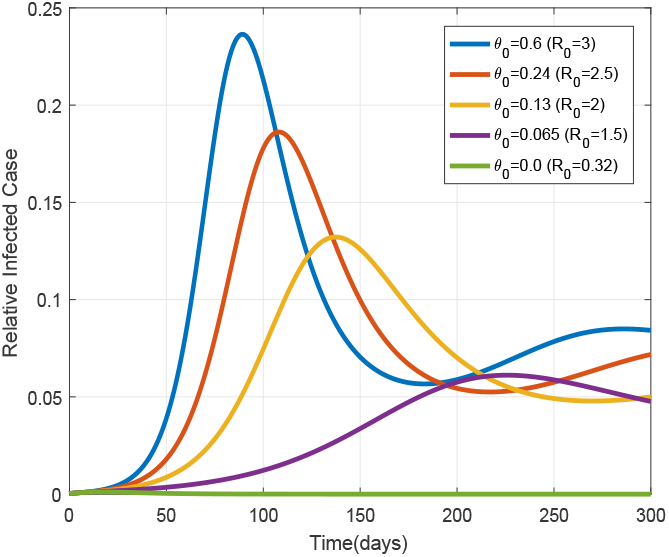
Impacts of *θ*_0_ on *w*(*t*).

## Conclusions

In this paper, a researcher investigated the dynamics of the COVID-19 spreading with control measure. An *SHEIQRD* Corona pandemic model with public health intervention has been presented, and analyzed theoretically as will as numerically. The theoretical analysis of the model are done. An essential epidemiological parameter value *ℛ*_0_ is derived by using the next generation matrix approach. Furthermore, we have shown that the disease free equilibrium globally asymptotically stable if *ℛ*_0_ ≤ 1 and unstable otherwise. For the case where *ℛ*_0_ > 1, the exists a unique endemic equilibrium *E*^*\**^, which is locally asymptotically stable. The sensitivity analysis of basic reproductive number was conducted. Its result suggested that transmission rate *β*, isolation rates *η* and *α*, stay at home rate *θ* and stay at home return rate *θ*_0_ are the basic control parameter of the model.

Numerical simulations are conducted aim to support theoretical analysis and shows the significance of public health intervention to containment these pandemics. The general dynamics of the model with time is illustrated that the disease is die out when *ℛ*_0_ ≤ 1 (see figure 10), but its persists in the community whenever *ℛ*_0_ > 1 (see figure 11). Moreover, socioeconomically crisis caused by these pandemic can be minimized and eliminated when we implemented appropriate control measure.

Also of importance in mitigation of the pandemics are reduced a transmission rate *β* (see figure 12) and the stay at home return rate *θ*_0_, an efficient identification and isolation of exposed and infected individual with rate of *η* and *α* (see figures 13, 14) respectively, and enhance the ability of stay at home rate *θ* (see figure 15). Finally, robust public health intervention end the current pandemic and minimizing crisis caused by these outbreak.

## Data Availability

I declare all data I used are available.

